# Viral load of SARS-CoV-2 Omicron BA.5 is lower than that of BA.2 despite the higher infectivity of BA.5

**DOI:** 10.1101/2022.10.25.22281427

**Authors:** Yuna Takatsuki, Yuta Takahashi, Jun Nakajima, Yumi Iwasaki, Katsutoshi Nagano, Chihiro Tani-Sassa, Sonoka Yuasa, Saki Kanehira, Kazunari Sonobe, Yoko Nukui, Hiroaki Takeuchi, Kousuke Tanimoto, Yukie Tanaka, Akinori Kimura, Naoya Ichimura, Shuji Tohda

## Abstract

Sublineage BA.5 of the SARS-CoV-2 Omicron variant rapidly spread and replaced BA.2 in July 2022 in Tokyo. A high viral load can be a possible cause of high transmissibility. Therefore, the copy numbers of SARS-CoV-2 in nasopharyngeal swab samples obtained from all patients visiting the hospital where this research was conducted were measured using quantitative polymerase chain reaction (qPCR). Viral genotypes were determined using PCR-based melting curve analysis. Next, whole-genome sequencing was performed using approximately one-fifth of the samples to verify the viral genotypes determined using PCR. Then, the copy numbers of the BA.1, BA.2, and BA.5 cases were compared. Contrary to expectations, the copy numbers of the BA.5 cases (median 4.7 × 10^4^ copies/μL, *n* = 290) were significantly (*p* = 0.001) lower than those of BA.2 cases (median 1.1 × 10^5^ copies/μL, *n* = 184). There was no significant difference between the BA.5 and BA.1 cases (median, 3.1 × 10^4^ copies/μL; *n* = 215). The results presented here suggest that the increased infectivity of BA.5 is not caused by higher viral loads, but presumably by other factors such as increased affinity to human cell receptors or immune escape due to its L452R mutation.

## 1. INTRODUCTION

The number of coronavirus disease 2019 (COVID-19) patients infected with the sublineage BA.1 of the Omicron variant of SARS-CoV-2 rapidly increased in Tokyo in January 2022. This spread of BA.1 was replaced by BA.2 in April 2022, after which the number of patients decreased gradually. However, the number of patients infected with the sublineage BA.5 abruptly increased, and BA.5 consequently replaced BA.2 by July 2022. This phenomenon was observed globally^1,2^ but its cause is currently unexplored. Therefore, examining the cause of the higher transmissibility of BA.5 relative to BA.1 and BA.2 remains crucial.

As a core hospital, the Tokyo Medical and Dental University (TMDU) Hospital has been mainly taking care of moderate or severe COVID-19 patients from all over Tokyo. This enabled the testing of the exact copy number and variant types of SARS-CoV-2 in nasopharyngeal swab samples from these patients who visited the hospital. The authors’ previous study has reported that there was no significant difference in the copy numbers between the Delta variant cases and Omicron variant cases, the latter of which mainly involved BA.1, as well as BA.2 to a certain extent^3^.

A possible mechanism for the high transmissibility of BA.5 may be a result of higher viral loads; therefore, to test this hypothesis, the copy numbers of viruses in the samples of patients infected with BA.1, BA.2, and BA.5 were compared.

## 2. MATERIALS AND METHODS

### 2.1 Patients and samples

A total of 774 COVID-19 patients who visited the TMDU hospital from February 1, 2021 to August 31, 2022, were diagnosed via polymerase chain reaction (PCR) tests. Nasopharyngeal swab samples were collected during a patient’s first outpatient visit or admission. The swabs were immersed in test tubes containing 1 mL of phosphate-buffered saline containing 1% dithiothreitol. Only the first sample obtained from each patient was used in this study. The Medical Research Ethics Committee of Tokyo Medical and Dental University gave ethical approval for this work (approval number: M2020-004). This study was performed in accordance with the 1964 Declaration of Helsinki.

### 2.2 Quantitative PCR test

The copy numbers of SARS-CoV-2 in the samples were measured using one-step reverse transcription-quantitative PCR (RT-qPCR) using the 2019 Novel Coronavirus Detection Kit (Shimadzu Corp., Kyoto, Japan) without RNA purification. The copy number per 1 μL of sample was determined by applying threshold cycle numbers of each sample to the calibration line of the standard samples of known concentration using the QuantStudio 5 Dx Real-Time PCR System (Thermo Fisher Scientific, Waltham, MA, USA).

### 2.3 Variant screening PCR test

The variants and their sublineage types were determined via melting curve analysis of the PCR products. RNA was extracted from swab samples using the EZ1 Virus Mini Kit v2.0 with EZ1 Advanced XL (QIAGEN, Venlo, The Netherlands). Reverse transcription-PCR was performed using primers and fluorescently labeled probes for N501Y, E484K, L452R, and S371L/S373P provided in the VirSNiP SARS-CoV-2 kits (TIB Molbiol; Berlin, Germany) alongside the LightCycler Multiplex RNA Virus Master (Roche Molecular Systems, Basel, Switzerland). Melting curve analysis was performed, and the variant and sublineage types were determined based on the resulting melting temperatures (T_m_).

### 2.4 Whole-genome sequencing (WGS) analysis

WGS was also performed using approximately one-fifth of the samples to verify the variant and sublineage types determined using PCR. Libraries prepared using the QIAseq SARS-CoV-2 Primer Panel Kit (QIAGEN) were sequenced on the MiSeq platform (Illumina, San Diego, CA, USA). Variant calling was performed using CLC Genomics Workbench software (QIAGEN).

### 2.5 Statistical analysis

The differences in copy number among sublineage cases were evaluated using the Wilcoxon rank-sum test and Steel–Dwass test for multiple comparisons.

## 3. RESULTS

A total of 774 samples were analyzed. The variant and sublineage types of 85 samples could not be determined because the variant screening PCR did not generate PCR products, mainly due to the very small copy numbers measured using quantitative PCR. S371L/S373P probe was used in the study, which could distinguish between BA.1 (371L/373P; T_m_: 62 °C) and BA.2/BA.4/BA.5 (371F/373P; T_m_: 53.5 °C), as well as the variants other than Omicron (371S/373S; T_m_: 45 °C), in the PCR samples. The types of all the samples that generated PCR products were determined to be of Omicron variants. Alpha variants (501Y, 484E, 452L, and 371S/373S) and Delta variants (501N, 484E, 452R, and 371S/373S) were not detected in the samples from this study period.

Specifically, 215 samples were carrying 501Y, 484A, 452L, and 371L/373P, designated BA.1, while 184 samples were carrying 501Y, 484A, 452L (T_m_ of 48 °C using L452R probe), and 371F/373P mutations, classified as BA.2. These BA.2 sample groups included six BA.2.12.1 samples with 501Y, 484A, 452Q (Tm 46 °C by L452R probe), and 371F/373P mutations and ten unnamed BA.2 samples with 501Y, 484A, 452M (T_m_ of 44 °C using L452R probe), and 371F/373P mutations.

Additionally, the variant PCR performed in this study identified 290 samples that were positive for 501Y, 484A, 452R, and 371F/373P mutations, and thus were classified as BA.5. Sublineage BA.5 viruses also carry an F486V mutation^1^, and this mutation consequently shifts the T_m_ of E484K probe to 37 °C, whereas the T_m_ of BA.2 samples with 484A and 486F remains at 40 °C. However, the variant PCR used in this study could not distinguish between BA.4 and BA.5 as both sublineages carry 501Y, 486V, 484A, 452R, and 371F/373P mutations.

The WGS analysis was performed on approximately one-fifth of the samples, and the genotyping results were consistent with those obtained using PCR. Considering that WGS analysis did not detect the BA.4 sublineage at all, the samples with 501Y, 486V, 484A, 452R, and 371F/373P were regarded as BA.5 in the following analysis although the presence of BA.4 sublineage in the samples remained a possibility. The WGS analysis did not find BA2.75^4^ in this study, either.

The sequential transition of BA.1, BA.2, and BA.5 are shown in Figure 1. Untyped samples were excluded from Figure 1 and the following analysis. The spread of BA.1 was replaced by BA.2 in April 2022, but the spread of BA.5 abruptly increased in the following July. The profiles of the 689 patients infected with BA.1, BA.2, and BA.5 are shown in Table 1. The distribution of viral copy numbers in swab-soaked samples from the three groups is shown in Figure 2.

**TABLE 1.** Profiles of patients infected with BA.1, BA.2, and BA.5 Omicron variants

| Sublineage type | BA.1 | BA.2 | BA.5 |
| --- | --- | --- | --- |
| Case numbers | 215 | 184 | 290 |
| Age (years) mean ± S.D. | 58 ± 23 | 51 ± 24 | 58 ± 24 |
| Viral load (copies/μL) median | 3.1 $\times 10^4$ | 1.1 $\times 10^5$ | 4.7 $\times 10^4$ |
| S.D.; standard deviation |  |  |  |

**FIGURE 1.**
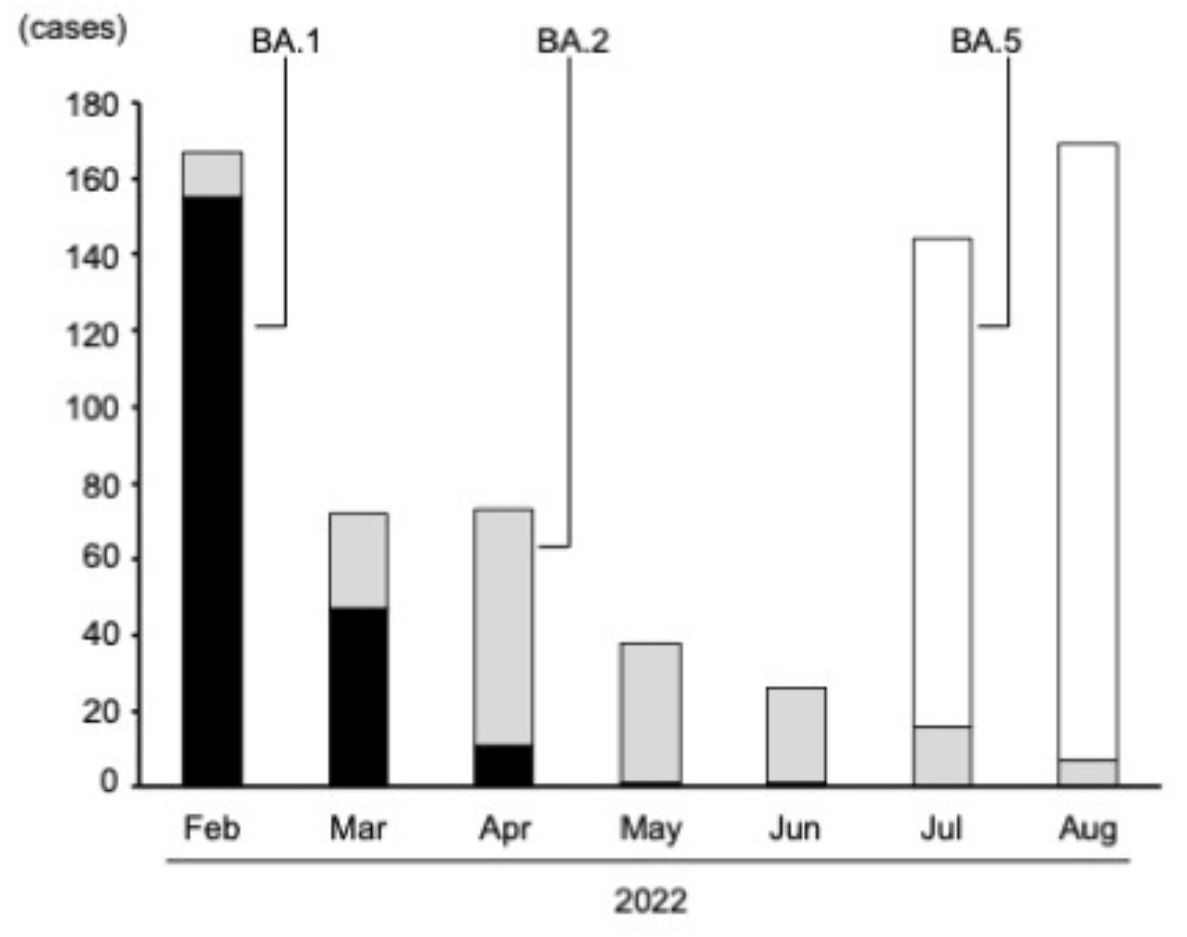
Sequential transition of BA.1, BA.2, and BA.5 Omicron variant cases determined using polymerase chain reaction (PCR)-based analysis.

**FIGURE 2.**
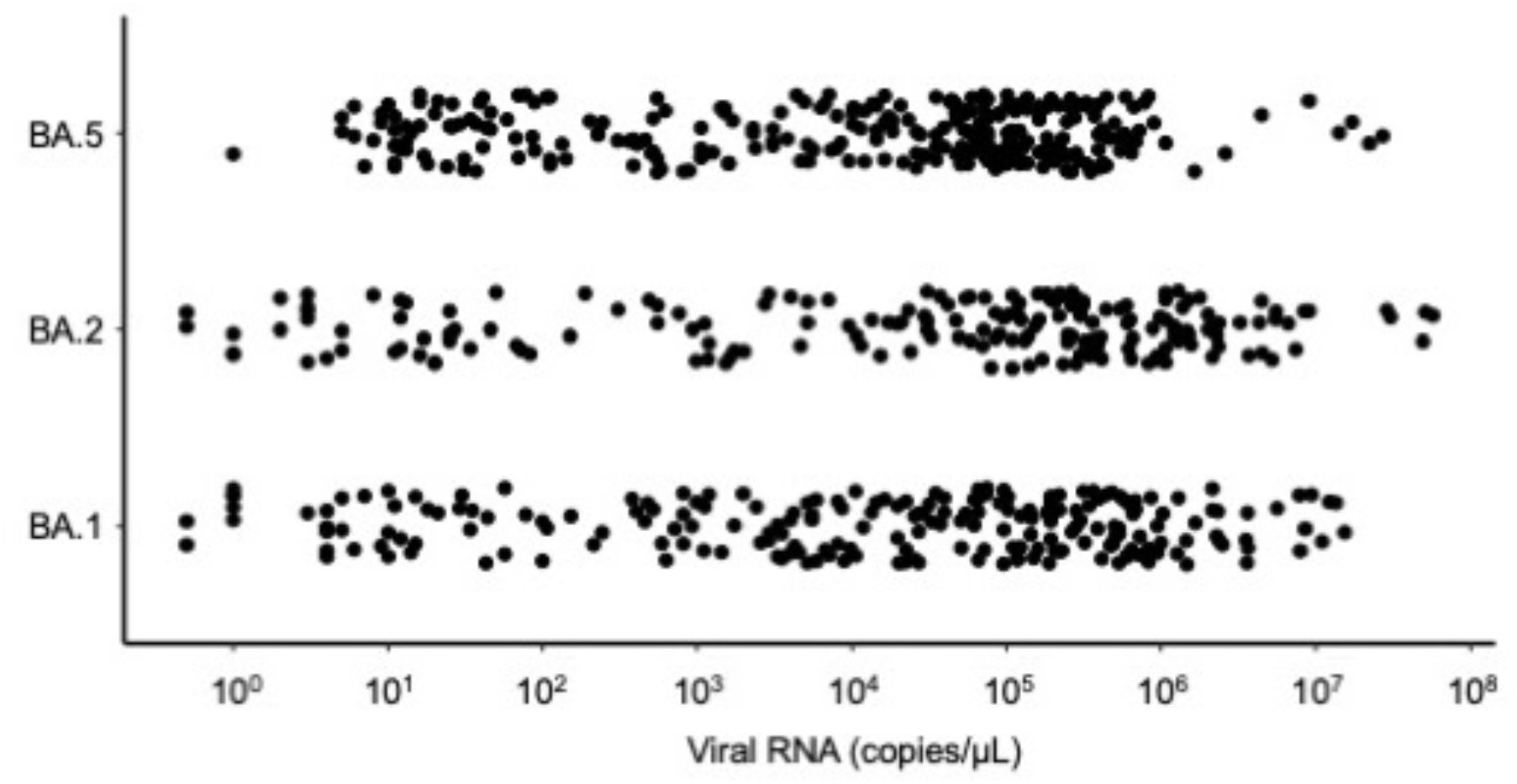
Distribution of the viral copy numbers of BA.1, BA.2, and BA.5 in swab-soaked samples measured using reverse transcription-quantitative polymerase chain reaction (RT-qPCR).

Contrary to expectations, the copy numbers in BA.5 cases (median 4.7 × 10^4^ copies/μL, *n* = 290) were significantly (*p* = 0.001) lower than those in BA.2 cases (median 1.1 × 10^5^ copies/μL, *n* = 184), although the distribution of copy numbers of each sublineage was widely dispersed as shown in Figure 2. There was no significant difference between the BA.5 and BA.1 cases (*p* = 0.52) or between the BA.2 and BA.1 cases (*p* = 0.11).

## 4. DISCUSSION

The Omicron BA.5 sublineage spread more rapidly than BA.1 and BA.2 and eventually replaced BA.2. This suggests that the BA.5 viruses exhibit a high transmissibility. It is surmised that a higher viral load could be one of the possible causes of higher transmissibility. However, contrary to expectations, the copy numbers observed in the BA.5 cases were significantly lower than those seen in the BA.2 cases although they were widely dispersed in both cases. This finding suggests that the cause of the high transmissibility of BA.5 is not due to its high viral load, but due to some other undetermined factors, which may involve an increased affinity to ACE2 receptor of human cells^5^ or a novel capacity for immune escape due to its L452R mutation^6^.

As reported previously by the authors^3^, the median copy number of Delta variant samples from July 2021 to January 2022 was 1.5 × 10^5^ copies/μL (*n* = 290). The median copy number of BA.5 in this study was significantly lower than that of Delta cases in a previous study (*p* < 0.0001).

It should be noted that the results presented here are biased because the TMDU hospital mainly specializes in the treatment of moderate or severe COVID-19 patients. Therefore, the results do not reflect all COVID-19 cases in Tokyo, which include cases that do not need treatment. However, this would not affect the comparison of copy numbers among the three sublineages, because the role of the TMDU hospital was the same throughout the study period. In fact, the sequential transition in the numbers of cases associated with each sublineage shown in Figure 1 was almost similar to that reported by the Tokyo Metropolitan Institute of Public Health^7^.

The spread of SARS-CoV-2 is influenced by the vaccination rate. A report by the Bureau of Social Welfare and Public Health of the Tokyo Metropolitan Government^8^ showed that the rate of Tokyo residents who were vaccinated three times was 4% on February 1, 53% on May 1, 60% on July 1, and 63% on August 31. The increase in the rate from February to May may be one of the reasons why the number of patients decreased during this period. The lagging vaccination rate after May could have led to the spread of BA.5 in July.

Indeed, several limitations are present in this study. As discussed above, these data may have been affected by the selection bias present in the cohort of patients involved in the analysis. Most patients infected with the Omicron variant are either asymptomatic or show mild symptoms. The proportion of patients with moderate or severe symptoms was higher in this study than in the general Tokyo population.

This study may be the first to compare copy numbers of Omicron sublineage cases, and the results suggest that the higher infectivity of Omicron BA.5 is not primarily caused by a higher viral load when compared to the viral load exhibited by previous variants and sublineages. More studies will be needed to clarify the precise mechanism of high infectivity of SARS-CoV-2 variants, which is an important aspect to understand in order to control the spread of COVID-19.

## Data Availability

All data produced in the present study are available upon reasonable request to the authors

## ACKNOWLEDGEMENTS

WGS was supported by the grant JPMJCR20H2 from JST-CREST and 20nk0101612h0901 from the Japan Agency for Medical Research and Development. We would like to thank all the staff involved in the COVID-19 treatment at the TMDU hospital.

## DATA AVAILABILITY STATEMENT

The data and RT-PCR protocol are available from the corresponding authors upon request.

## AUTHOR CONTRIBUTIONS

Study conception and design: Chihiro Tani-Sassa, Katsutoshi Nagano, Yumi Iwasaki, Naoya Ichimura, Yoko Nukui, and Shuji Tohda. Sample analysis: Yuna Takatsuki, Yuta Takahashi, Jun Nakajima, Sonoka Yuasa, Saki Kanehira, and Kazunari Sonobe. WGS: Hiroaki Takeuchi, Kousuke Tanimoto, Yukie Tanaka, and Akinori Kimura. Data analysis: Naoya Ichimura and Shuji Tohda. Writing: Shuji Tohda.

